# Low circulating adropin concentrations predict increased risk of cognitive decline in community-dwelling older adults

**DOI:** 10.1101/2023.03.24.23285476

**Authors:** Geetika Aggarwal, John E. Morley, Bruno Vellas, Andrew D. Nguyen, Andrew A. Butler, the MAPT/DSA Group

**Affiliations:** Henry and Amelia Nasrallah Center for Neuroscience, Saint Louis University, St. Louis, MO, USA; Division of Geriatric Medicine, Department of Internal Medicine, Saint Louis University School of Medicine, St. Louis, MO, USA; Department of Pharmacology & Physiology, Saint Louis University School of Medicine, St. Louis, MO, USA; Gérontopôle de Toulouse, Institut du Vieillissement, Centre Hospitalo-Universitaire de Toulouse, 37 allées Jules Guesdes, 31000, Toulouse, France

## Abstract

The secreted peptide adropin is highly expressed in human brain tissues and correlates with RNA and proteomic risk indicators for dementia. Here we report that plasma adropin concentrations predict risk for cognitive decline in the Multidomain Alzheimer Preventive Trial (ClinicalTrials.gov Identifier, NCT00672685; mean age 75.8y, SD=4.5y, 60.2% female, n=452). Cognitive ability was evaluated using a composite cognitive score (CCS) that assessed four domains: memory, language, executive function, and orientation. Relationships between plasma adropin concentrations and changes in CCS (ΔCCS) were examined using Cox Proportional Hazards Regression, or by grouping into tertiles ranked low to high by adropin values and controlling for age, time between baseline and final visits, baseline CCS, and other risk factors (e.g., education, medication, APOE4 status). Risk of cognitive decline (defined as a ΔCCS of -0.3 or more) decreased with increasing plasma adropin concentrations (hazard ratio = 0.873, 95%CI 0.780-0.977, P=0.018). Between adropin tertiles, ΔCCS was significantly different (P=0.01; estimated marginal mean±SE for the 1^st^-to 3^rd^-tertile, -0.317±0.064; -0.275±0.063; -0.042±0.071; n=133,146, and 130, respectively; P<0.05 for 1^st^ vs. 2^nd^ and 3^rd^ adropin tertiles). Normalized plasma Aβ_42/40_ ratio and plasma neurofilament light chain, indicators of neurodegeneration, were significantly different between adropin tertile. These differences were consistent with reduced risk of cognitive decline with higher plasma adropin levels. Overall, these results suggest cognitive decline is reduced in community-dwelling older adults with higher circulating adropin levels. Further studies are needed to determine the underlying causes of the relationship and whether increasing adropin levels can delay cognitive decline.

## INTRODUCTION

Aging is the most significant risk factor for cognitive decline and dementia, a general term used to describe disorders adversely affecting intellectual ability (for e.g., memory impairment, aphasia, confusion, disorientation) (1). Gains in life expectancy are markedly increasing the number of individuals requiring treatment for dementia (2, 3). Blood-based biomarkers have been developed that identify at-risk individuals experiencing the early stages of Alzheimer’s disease (AD), the most common form of dementia (4-7). Biomarkers reflecting brain amyloid accumulation (plasma Aβ_42/40_ ratio), pathologic tau protein (p-tau231, p-tau181, p-tau217) and neurodegeneration (neurofilament light chain (NfL)) have advanced the diagnosis of AD and other neurodegenerative diseases (4-7). However, treatment options remain limited. Most of the medications target the symptoms of dementia, while the efficacy of the recently approved medications that target the underlying biology is controversial (1, 8). An urgent need continues to exist for identifying modifiable, easily measurable risk factors and new treatment strategies.

Adropin is a short 76 amino acid peptide encoded by the Energy Homeostasis Associated (ENHO) gene (9). While originally described as a secreted peptide (9), further study suggests adropin^1-76^ might also reside in the plasma membrane (10). This conclusion is supported by data from AlphaFold’s transmembrane protein structure database (11) and the HMMTOP algorithm (12). Adropin immunoreactivity is nevertheless observed in blood specimens and in media of cultured cells (9), suggesting release of some part of the adropin peptide into the interstitial space. A synthetic peptide derived from the putative extracellular domain (adropin^34-76^) induces biological responses in cultured cells and rodent models that parallel changes observed with transgenic over expression or deletion of the full-length protein (13, 14).

Experiments using mouse models suggest adropin regulates physiological processes relevant to healthy neurological aging. Insulin signaling and glycemic control are important drivers of aging processes and longevity (15). In mice, adropin acts as an insulin sensitizer and directly regulates glucose metabolism (16-22). Adropin actions also preserve arterial elasticity in the context of aging and type 2 diabetes (23-26). Finally, an emerging literature indicates that adropin acts on the cerebral vasculature to preserve neurological functions during cerebral ischemia (27-29).

The role of adropin in human aging is less clear. Cross-sectional data indicate relationships with risk indicators for diabetes and vascular disease (13, 14, 24, 26). However, expression profiling supports a more direct relationship between brain adropin expression and aging-related neurological conditions in humans. Expression of the transcript encoding adropin is higher by orders of magnitude in the central nervous system relative to other tissues, suggesting that it functions primarily as a neuropeptide (10, 30, 31). Adropin expression in the brain correlates with proteomic and transcriptomic signatures for risk of cognitive decline (30). Positive correlations with transcriptomic signatures of mitochondrial and synaptic functions suggest adropin enhances synaptic plasticity and glucose utilization (30). On the other hand, adropin expression also correlates positively with markers of Aβ accumulation and Tau pathology (30). Experiments using mouse models indicate that increased adropin activity preserves cognitive ability in the context of aging, metabolic stressors associated with obesity, or cerebral ischemia (28, 30, 32, 33).

The relationship between circulating adropin levels and aging-related cognitive decline has however not been reported. Here we report an investigation of the relationships between plasma adropin concentrations and cognitive decline in community dwelling older adults.

## METHODS

### Study population

The study participants were from the Multidomain Alzheimer Preventive Trial (MAPT, ClinicalTrials.gov NCT00672685), a randomized controlled trial that assessed the impact of nutritional supplement (omega-3 fatty acid) alone or in combination with a multidomain intervention. The original study examined 1679 dementia-free older adults aged ≥ 70 years recruited with any of the following criteria: expressing spontaneous memory complaint; having limitation in at least one instrumental activity of daily, and slow gait speed (<0.8 m/s). Participants were excluded if any of the following criteria was met: a mini-mental state exam (MMSE) score ≤24, a diagnosis of dementia, exhibiting difficulties in performing the basic activities of daily living, and already taking polyunsaturated fatty acid supplementation. The MAPT tested multidomain interventions (physical activity, nutritional counselling, and cognitive training) and omega-3 supplementation, combined or alone, against placebo among older adults and examined changes in cognitive functions over a 3-year period (34). Participants were observed for two additional years, without receiving any intervention. The MAPT was approved by the ethics committee in Toulouse (CPP SOOM II). Written consent forms were obtained from all participants. All research was performed in accordance with relevant guidelines/regulations.

### Calculation of the Compositive Cognitive Score (CCS)

Participants completed a comprehensive assessment of four domains: memory (free and total recall of the Free and Cued Selective Reminding Test [FCSRT]), language (the Category Naming Test), executive function (the DSST-WAISR) and orientation (ten MMSE orientation items) (35). There was a total of 11 visits (V1-V11) during the study. Cognitive testing occurred on the first two visits, and then annually on the odd-number visits (V1, V2, V3, V5, V7, V9, and V11). The plasma used to measure adropin concentrations was collected on V3 (n=419) and V5 (n=33).

CCS were calculated using the average of the Z-score for each domain using values at V1 as the reference point. For calculation of the Z scores, data collected at V1 was used to calculate the initial mean (*μ*^*B*^) and SD (*σ*^*B*^) for each specific test result (FCSRT, Category Naming Test, DSST-WAISR, MMSE orientation items). These values were used to calculate a Z score for the data collected at baseline and subsequent visits: Z = (*x* ^*V*^ *-μ* ^*B*^ ) ÷ *σ* ^*B*^) where *x*^*V*^ is the test result score for each test on each visit (V1, V2, etc.), *μ* ^*B*^ is the mean of the test scores for each specific test at baseline, and *σ*^*B*^ is the SD of the test scores for each specific test at baseline. The CCS was then calculated by taking the average Z score for each test result at each time point.

Two approaches for comparing adropin values with ΔCCS are presented. Plasma used for the study came mostly from V3 (n=419), which was 1 year after V1, and from V5 (n=33), which occurred 2 years after V1. We initially compared plasma adropin concentrations collected at V3/V5 with ΔCCS calculated over the full duration of the study, using CCS at V1 as the baseline. The advantage of this approach was in maximizing the time during which changes in CCS could happen (up to 5 years). The disadvantage is that the actual plasma adropin concentrations at baseline are not known, and any correlations observed are between adropin value collected at an arbitrary mid-point in the study. To address this weakness, we also calculated ΔCCS calculated using CCS values at V3 and V5 as baseline. This approach allowed us to compare plasma adropin concentrations with subsequent changes in CCS using the same time point as baseline. This comparison provides a more clearly defined test of whether plasma adropin concentrations test at any given age in this cohort predict risk of subsequent decline. However, this approach reduced the effective duration between the first and final cognitive tests from 5 years to 4 years for the V3 timepoint, and to 3 years for the V5 timepoint.

For each approach, changes in CCS over time (ΔCCS) were calculated by subtracting the final score from the baseline score. To control for variability in time between the baseline and final visit, we used a regression approach with years between the baseline and final visit included as a covariate.

This study compared CCS in participants without dementia. For this group of 452 participants, 37 were diagnosed with dementia. Data from the visit during which these participants were diagnosed with dementia was excluded from the analysis.

### Measurement of plasma variables

Most plasma samples used to measure adropin concentrations were collected at V3 (n=419), with a smaller number collected at V5 (n=33). This selection was based on the availability of plasma samples. Cross-sectional comparisons of plasma adropin concentrations with other plasma variables used plasma collected on V3. Cross-sectional comparisons of plasma adropin concentrations with physiological measures (BMI, heart rate, blood pressure) used data collected on V3 and V5.

Plasma adropin concentrations were measured using an enzyme immunoassay kit from Phoenix Pharmaceuticals, Inc. (cat. no. EK-032-35) following the manufacturer’s protocol. Assay sensitivity reported by the manufacturer is 0.3 ng/ml, with a linear range from 0.3 to 8.2 ng/ml. Pilot experiments determined a 1:5 dilution of serum produced values within the linear range. Values >8.2 ng/ml were described as “high” and arbitrarily assigned a value of 8.5 ng/ml (19 out of 452 samples). The assay was performed in duplicates; values with a CV of >20% were discarded. Plate controls included ‘in-house’ human plasma and the controls provided by the assay manufacturer. The %CV for the plate controls were 11% and 14%, respectively.

The methods used and a description of the results from the measurement of other plasma variables reported here have been previously described (34, 36-39).

### Statistical analysis

Data were collated and managed in Microsoft Excel prior to import into SPSS vers. 28.0.1.0 (IBM). Relationships between plasma adropin concentration and declining CCS were initially modelled using cox binomial regression. Cognitive decline for this study was defined as a change in CCS (ΔCCS) of -0.3 or greater. Covariates used for the analysis include the number of years between V1 and the final visit used to calculate ΔCCS, sex (categorical) and APOE status (categorical, APOE ε4 positive or negative), BMI at V1, medications, and years of education. Cognitive decline was also compared between participants with lower than normal or higher than normal adropin. For this approach, participants were separated into adropin tertiles using the 33^rd^ and 67^th^ percentiles of ranked data. As plasma adropin concentrations were significantly different between sex; assignment was performed separately within sex. The 1^st^, 2^nd^, and 3^rd^ tertile thus corresponded with participants with low, ‘normal’, or high levels for each sex.

A numerical score was used to control for education (1=no diploma or less than primary school certificate, 2=primary school certificate, 3=secondary education, 4=high school diploma, 5=university level). To control for medication, study participants received scores of 0 (not prescribed) or 1 for drugs targeting the nervous system (analgesics, anesthetics, anti-epileptics, psychoanaleptics, psycholeptics, anti-parkinson drugs), lipid lowering agents (statins, fibrates, bile acid sequestrants, cholesterol absorption inhibitors), diabetes drugs (alpha glucosidase inhibitors, sulfonamides, thiazolidinediones, DPP-4 inhibitors, biguanides, insulin analogs), drugs targeting the cardiovascular system (ACE inhibitors, ARB blockers, anti-adrenergic drugs), drugs for blood disorders (antithrombotic and antianemic agents), corticosteroids, anti-inflammatory and antirheumatic drugs, and thyroid medications. The participants received possibles score between 0 (no medication) and 8.

Tertiling was also used for comparisons of morphometry and plasma data between adropin tertiles. Data with equal distributions around the mean, or which passed the test following transformation (Log_10_), were analyzed using ANCOVA. The covariates used for each analysis are indicated; post hoc tests between groups used Bonferroni to adjust for multiple comparisons. Data failing test for homogeneity of variance were compared using a nonparametric test (Quade Nonparametric Analysis of Covariance) with covariates applied as indicated. Adjusted data are presented as estimated marginal means and std. error.

## RESULTS

Plasma adropin concentrations did not correlate with age, BMI, or educational status, and there were no significance for these parameters between adropin tertiles (**Table 1**). Plasma adropin concentrations and CCS were significantly different between sex (females>males, **Table 1**).

**Table 1.** Plasma adropin concentrations, CCS, and morphometric data for subjects grouped into adropin tertiles. Plasma adropin concentrations and age are mean and SD. Baseline CCS are estimated marginal means and SE adjusted for age, medication, and years of education and are measurements recorded on the visit plasma samples were collected (V3, V5). Body mass index (BMI) are estimated marginal mean adjusted for age. Sample sizes are indicated in brackets. P values for covariates less than 0.05 are shown in *italics*.

| Metric | Sex | Adropin tertile |  |  |  | P value |
| --- | --- | --- | --- | --- | --- | --- |
|  |  | (ALL) | 1st | 2nd | 3rd |  |
| Plasma Adropin (ng/ml) | F | 4.21±1.70<br>(272) | 2.43±0.77<br>(90) | 4.12±0.39<br>(91) | 6.06±1.16<br>(91) | Sex<br>P<0.01 |
|  | M | 3.73±1.80<br>(180) | 2.05±0.47<br>(60) | 3.38±0.38<br>(61) | 5.80±1.51<br>(59) |  |
|  | ALL | 4.02±1.76<br>(452) | 2.34±1.01<br>(150) | 3.83±0.53<br>(152) | 5.94±1.40<br>(150) | Tertile<br>P<0.01 |
| CCS | F | +0.180±0.042<br>(265) | +0.195±0.072<br>(88) | +0.206±0.071<br>(91) | +0.139±0.073<br>(86) | *Education, Age*<br>*P<0.001* |
|  | M | -0.055±0.051<br>(174) | -0.015±0.090<br>(56) | -0.130±0.087<br>(61) | -0.021±0.091<br>(57) |  |
|  | ALL | 0.062±0.033<br>(439) | 0.090±0.057<br>(144) | 0.038±0.056<br>(152) | 0.059±0.058<br>(143) | Sex<br>P<0.01 |
| Age, V1 (yr) | F | 75.6±4.4<br>(272) | 76.2±4.7<br>(90) | 75.0±4.2<br>(91) | 75.8±4.3<br>(91) |  |
|  | M | 76.0±4.7<br>(180) | 75.5±4.5<br>(60) | 75.9±4.6<br>(61) | 76.8±5.0<br>(59) |  |
|  | ALL | 75.8±4.5<br>(452) | 75.9±4.6<br>(150) | 75.3±4.3<br>(152) | 76.2±4.6<br>(150) |  |
| BMI (kg/m<sup>2</sup>) | F | 25.8±0.2<br>(271) | 25.7±0.4<br>(89) | 26.3±0.4<br>(91) | 25.5±0.4<br>(91) |  |
|  | M | 26.7±0.3<br>(179) | 27.0±0.5<br>(60) | 26.8±0.5<br>(60) | 26.3±0.5<br>(59) |  |
|  | ALL | 26.3±0.2<br>(450) | 26.4±0.3<br>(149) | 26.6±0.3<br>(151) | 25.9±0.3<br>(150) |  |
| **Education level (%)** |  |  |  |  |  |  |
| Less than primary |  | 5% | 6% | 4% | 4% |  |
| Primary |  | 20% | 20% | 22% | 19% |  |
| Secondary |  | 34% | 38% | 32% | 32% |  |
| High school diploma |  | 14% | 13% | 16% | 14% |  |
| University |  | 27% | 23% | 27% | 31% |  |
|  |  | (N=446) | (N=145) | (N=152) | (N=149) |  |

However, a cross-sectional comparison of plasma adropin concentrations indicate no correlations with CCS recorded on the visit used to collect plasma samples used for this study (V3, n=419; V5, n=33; ρ=0.046) (**Table 1**).

### Slower aging-related cognitive decline with higher plasma adropin concentrations

A decline in cognitive score of -0.3 or more in one calendar year has been reported to predict increased risk for a future dementia diagnosis (40). For this study, a decline in CCS of -0.3 or more during the 5 years of the MAPT study observed in 157 participants (35.5%) was used to define as a significant event. Simple modelling using Cox Proportional Hazards regression indicated plasma adropin concentrations predict risk of decline. Controlling for age at V1, the number of years between visits used to calculate ΔCCS, and stratification by APOE4 status (null, one or more copies) indicated declining risk with increasing plasma adropin concentrations (HR=0.898, 95% CI 0.808-0.998, P=0.045). Plasma concentrations of NfL and normalized plasma AB_42/40_ ratio are biomarkers of neuropathology and were significantly different between adropin tertiles (**Table 2**). APOE4 status also appeared to be lower (24% vs. 31%) in the 3^rd^ adropin tertile (**Table 2**). However, years of education and medication usage was similar between tertiles (**Tables 1, 3-5**). The addition of these and other potentially confounding variables to the model (BMI, sex) did not markedly change the result (HR=0.873, 95% CI 0.780-0.977, P=0.018).

**Table 2.** Plasma concentrations of markers of neurodegeneration (NfL, AB_42/40_). Plasma normalized Aβ_42/40_ ratio and NfL are presented as estimated marginal mean and SE adjusted for age. P values for covariates less than 0.05 are shown in *italics*. For APOE4 status, percent exhibiting at least one copy are shown.

| Metric | Sex | Adropin tertile |  |  |  | P value |
| --- | --- | --- | --- | --- | --- | --- |
|  |  | (ALL) | 1st | 2nd | 3rd |  |
| A$\beta_{42/40}$<br>(ratio) | F | 0.115±0.001<br>(251) | 0.114±0.002<br>(83) | 0.114±0.002<br>(85) | 0.116±0.002<br>(83) | Tertile<br>P<0.05 |
|  | M | 0.112±0.001<br>(171) | 0.110±0.001<br>(56) | 0.109±0.002<br>(61) | 0.118±0.002<sup>1</sup><br>(54) |  |
|  | ALL | 0.114±0.001<br>(422) | 0.112±0.002<br>(139) | 0.111±0.002<br>(146) | 0.117±0.002<sup>1</sup><br>(137) |  |
| NfL<br>(pg/mL) | F | 85.8±4.5<br>(248) | 80.0±7.7<br>(71) | 97.4±7.7<br>(87) | 79.9±7.7<br>(90) | *Age,*<br>*P=0.005*<br>*Sex X*<br>*Adropin*<br>*tertile,*<br>*P<0.05* |
|  | M | 87.9±5.5<br>(171) | 101.3±9.5<sup>2</sup><br>(53) | 84.4±9.4<br>(59) | 78.1±9.6<br>(59) |  |
|  | ALL | 86.9±3.5<br>(419) | 90.7±6.1<br>(124) | 90.9±6.1<br>(146) | 79.0±6.2<br>(149) |  |
| APOE4<br>(%ve) | F | 28%<br>(248) | 33%<br>(82) | 24%<br>(86) | 26%<br>(80) |  |
|  | M | 27%<br>(166) | 27%<br>(55) | 33%<br>(60) | 20%<br>(51) |  |
|  | ALL | 28%<br>(414) | 31%<br>(137) | 28%<br>(146) | 24%<br>(141) |  |

<sup>1</sup> P<0.05 vs. 1<sup>st</sup> and 2<sup>nd</sup> adropin tertiles (within row).
<sup>2</sup> P<0.05 vs. 2<sup>nd</sup> and 3<sup>rd</sup> adropin tertiles (within row), P<0.05 for the 1<sup>st</sup> adropin tertile between sex.

**Table 3.** Indices of cardiovascular function. Data shown are estimated marginal means and SE adjusted for age and BMI. The % taking medications for cardiovascular conditions (antihypertensives) during the first 3 years of the study P values for covariates less than 0.05 are shown in *italics*.

| Metric | Sex | Adropin tertile |  |  |  | P value |
| --- | --- | --- | --- | --- | --- | --- |
|  |  | (ALL) | 1st | 2nd | 3rd |  |
| Pulse rate (standing) | F | 75.4±0.7<br>(269) | 73.7±1.2<sup>2</sup><br>(89) | 75.0±1.2<br>(91) | 77.5±1.2<sup>1,2</sup><br>(89) | *BMI, P<0.05*<br>Sex, P<0.001<br>Tertile, P<0.05<br>Sex X tertile, P<0.05 |
|  | M | 71.4±0.9<br>(178) | 69.1±1.5<br>(59) | 74.9±1.5<sup>1</sup><br>(60) | 70.1±1.5<br>(59) |  |
|  | ALL | 73.4±0.5<br>(447) | 71.4±1.0<br>(148) | 75.0±0.9<sup>1</sup><br>(151) | 73.8±1.0<br>(148) |  |
| Pulse rate (prone) | F | 70.3±0.6<br>(270) | 67.6±1.1<br>(89) | 70.6±1.1<br>(91) | 72.8±1.1<sup>4,5</sup><br>(90) | *BMI, P<0.05*<br>Sex, P=0.005<br>Tertile, P<0.001<br>Sex X Tertile, P=0.052 |
|  | M | 67.5±0.8<br>(178) | 64.3±1.4<br>(59) | 70.9±1.3<sup>5</sup><br>(60) | 67.2±1.4<br>(59) |  |
|  | ALL | 68.9±0.5<br>(448) | 65.9±0.9<sup>3</sup><br>(148) | 70.7±0.9<br>(151) | 70.0±0.9<br>(149) |  |
| Systolic blood pressure (standing) | F | 134.6±1.2<br>(269) | 131.2±2.2<br>(89) | 136.7±2.1<br>(91) | 135.9±2.2<br>(89) | *Age, P=0.001*<br>*BMI P=0.001* |
|  | M | 137.4±1.5<br>(178) | 138.4±2.7<br>(59) | 136.5±2.6<br>(60) | 137.4±2.7<br>(59) |  |
|  | ALL | 136.0±1.0<br>(447) | 134.8±1.7<br>(148) | 136.6±1.7<br>(151) | 136.6±1.7<br>(148) |  |
| Systolic blood pressure (Prone) | F | 137.0±1.1<br>(270) | 134.4±2.0<br>(89) | 138.5±2.0<br>(91) | 137.9±2.0<br>(90) | *Age, P=0.003*<br>*BMI P=0.002* |
|  | M | 139.1±1.4<br>(178) | 142.1±2.4<br>(59) | 138.7±2.4<br>(60) | 136.7±2.4<br>(59) |  |
|  | ALL | 138.0±0.9<br>(448) | 138.3±1.6<br>(148) | 138.6±1.6<br>(151) | 137.3±1.6<br>(149) |  |
| Diastolic blood pressure (standing) | F | 78.7±0.8<br>(269) | 75.9±1.3<br>(89) | 79.2±1.3<br>(91) | 81.2±1.3<sup>6</sup><br>(89) | *BMI, P<0.001*<br>Sex X tertile P=0.055 |
|  | M | 79.5±0.9<br>(178) | 79.2±1.6<br>(59) | 81.5±1.6<br>(60) | 77.8±1.6<br>(59) |  |
|  | ALL | 79.1±0.6<br>(447) | 77.5±1.1<br>(148) | 80.4±1.0<sup>1</sup><br>(151) | 79.5±1.1<br>(148) |  |
| Diastolic blood pressure (prone) | F | 77.4±0.7<br>(270) | 76.1±1.3<br>(89) | 76.9±1.3<br>(91) | 79.2±1.6<br>(90) | *BMI, P<0.001*<br>Sex X tertile P=0.056 |
|  | M | 78.0±0.9<br>(178) | 79.8±1.6<br>(59) | 78.1±1.6<br>(60) | 76.1±1.6<br>(59) |  |
|  | ALL | 77.7±0.6<br>(448) | 77.9±1.0<br>(148) | 77.5±1.0<br>(151) | 77.7±1.0<br>(149) |  |
| Cardiovascular medications (% prescribed) | ALL | 44.9% | 42.0% | 48.7% | 44.0% |  |

<sup>1</sup> P<0.05 vs. 1<sup>st</sup> adropin tertile within row (All), P<0.01 vs. 1<sup>st</sup> adropin tertile and <0.05 vs. 3<sup>rd</sup> adropin tertile within row (males) ; <sup>2</sup> P<0.001 (3<sup>rd</sup> tertile) or <0.05 (1<sup>st</sup> tertile) between sexes (within column); <sup>3</sup> P<0.001 vs. 2<sup>nd</sup>, 3<sup>rd</sup> adropin tertiles (within row); <sup>4</sup> P=0.001 between sexes (within column); <sup>5</sup> P<0.001 vs. 1<sup>st</sup> adropin tertile (within row), and <sup>6</sup> P=0.005 vs. 1<sup>st</sup> adropin tertile (within row)

Plotting ΔCCS as a function of age at V1 provided further indication that the acceleration of cognitive decline with aging was attenuated in the 3rd adropin tertile (**Fig. 1**). Cumulative hazard as a function of age at V1 was calculated controlling for APOE status, sex, BMI, normalized plasma AB_42/40_ ratio, education, and medications. As predicted, cumulate hazard increased as a function of age, and the increase was delayed for participants in the 3^rd^ adropin tertile (**Fig. 2A**). Comparing ΔCCS by 2-way ANCOVA (groups: sex, adropin tertile) also indicated delayed decline in the 3^rd^ adropin tertile (**Fig. 2B**). When adjusted for CCS and age at V1, years between first and final CCS measurement, years of education and medication, there was a strong trend for a difference between adropin tertiles (F_2,446_=2.931, P=0.054; estimated marginal mean±SE of ΔCCS for 1^st^, 2^nd^, and 3^rd^ adropin tertiles, -0.298±0.059, -0.253±0.057, and -0.104±0.059, P=0.065 between 1^st^ and 3^rd^ adropin tertiles). There was still a strong trend when other confounders (NfL, normalized plasma Aβ_42/40_ ratio, and BMI) were included as covariates (F_2,417_=2.720, P=0.067). There was no effect of sex in this analysis (data not shown).

**Figure 1.**
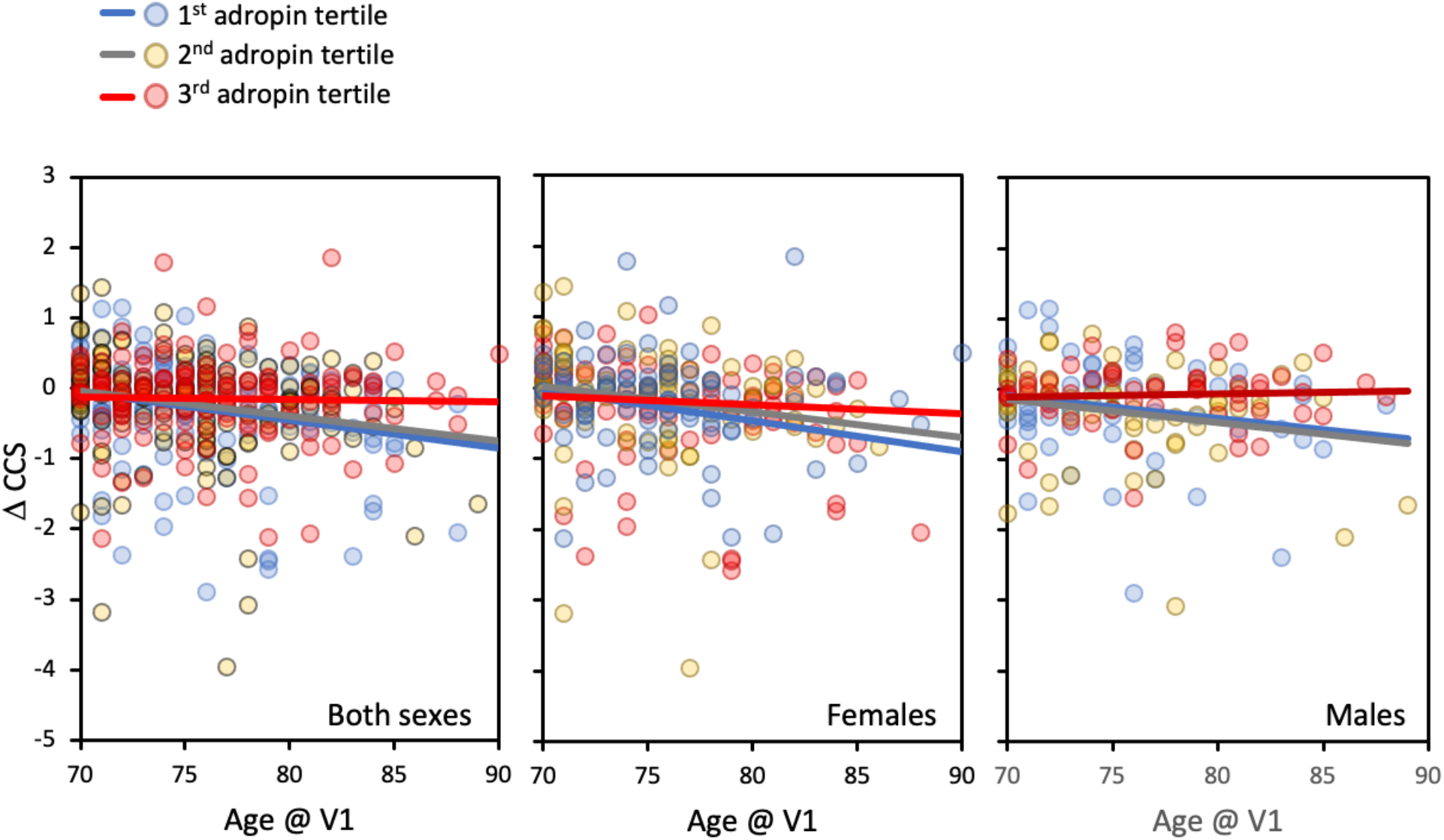
Scatterplots showing changes in composite cognitive score (ΔCCS) as a function of age on the first visit (V1). For these plots, ΔCCS was calculated using V1 as the baseline. The participants are divided into three groups (adropin tertiles) ranked low (1^st^) to high (3^rd^) For the 1^st^, 2nd, and 3rd adropin tertile, n=151, 152, and 149, respectively. The three panels show data for both sexes (left), females only (middle), and males only (right). The results from this analysis suggest that ΔCCS declines as a function of age for participants in the 1^st^ and 2^nd^ adropin tertiles, but not for participants in the 3^rd^ adropin tertile.

**Figure 2.**
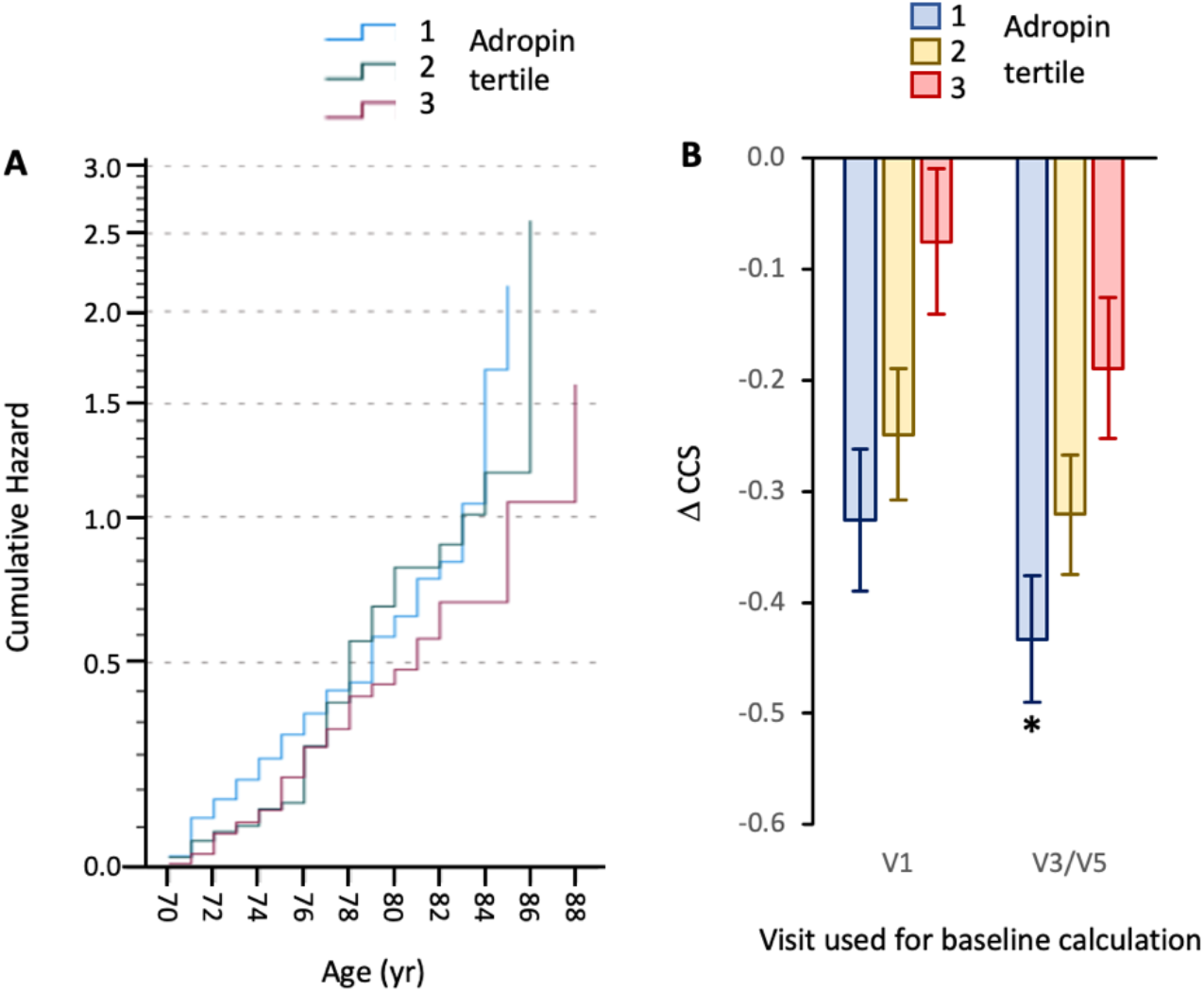
Cumulative hazard as a function of age (**A**) and comparing ΔCCS between adropin tertiles (**B**). (A) Cumulative hazard for exhibiting a decline of CCS of -0.3 or greater increases with age. The rate of increase is delayed for participants in the 3^rd^ adropin tertile relative to those in the 1^st^ or 2^nd^ adropin tertiles. (**B**) Estimated marginal means ± SEM for the ΔCCS for participant grouped into adropin tertiles. The data are adjusted for age, sex, years between baseline and final measurement, NfL, and Aβ_42/40_ ratio (37, 39), and APOE4 status. For the data using V1 as baseline, n=127,140, and 119, respectively; for data using V3/V5 as baseline, n=124,133, and 104, respectively. Significantly different from the 3^rd^ adropin tertile, * P<0.05.

The initial comparison used plasma adropin concentrations collected mid-study with changes in CCS over the 5 years of data collection. However, using CCS results collected at the same visits used for blood plasma collection (V3, V5) yielded similar outcomes. Changes in CCS from V3 or V5 adjusted for age at V1, years between visits used for baseline (V3 or V5) and final measurements of CCS, education, medical treatment, and baseline CCS values exhibited a strong trend for differences between adropin tertiles (F_2,413_=2.999, P=0.051; ΔCCS using V3/V5 as baseline for 1^st^, 2^nd^, and 3^rd^ adropin tertiles, -0.377±0.053; -0.330±0.053, or -0.191±0.057, n=141/145/127; P=0.054 between 1^st^ and 3^rd^ adropin tertiles). When controlling for other confounding variables, the differences between adropin tertiles were statistically significant (F_2,386_=4.043, P=0.018) (**Fig. 2B**). For the covariates used in the analysis, age, plasma Aβ_42/40_ ratio, and years between V3/V5 and final visit had significant effects (P<0.05), however there was still no significant effect of sex (data not shown).

The APOE4 allele is the strongest genetic risk factor identified for AD (41, 42). A 2-way ANCOVA used adropin tertile and APOE4 status as independent variables controlling for age, years between visits, plasma Aβ_42/40_ ratio, NfL, education, and medication indicated comparable outcomes between APOE genotypes (**Fig. 3A, B**). As predicted, there was a significant difference between adropin tertile when using either V1 (F_2,382_=4.705; P=0.010, **Fig. 3A**) or V3/V5 (F_2,381_=4.306, P=0.014, **Fig. 3B**) as baseline for calculating ΔCCS. While there was no interaction between APOE ε4 genotype and adropin tertile, in the post-hoc analysis significant differences between adropin tertile were only observed in the APOE4 allele group (**Fig. 3**).

**Figure 3.**
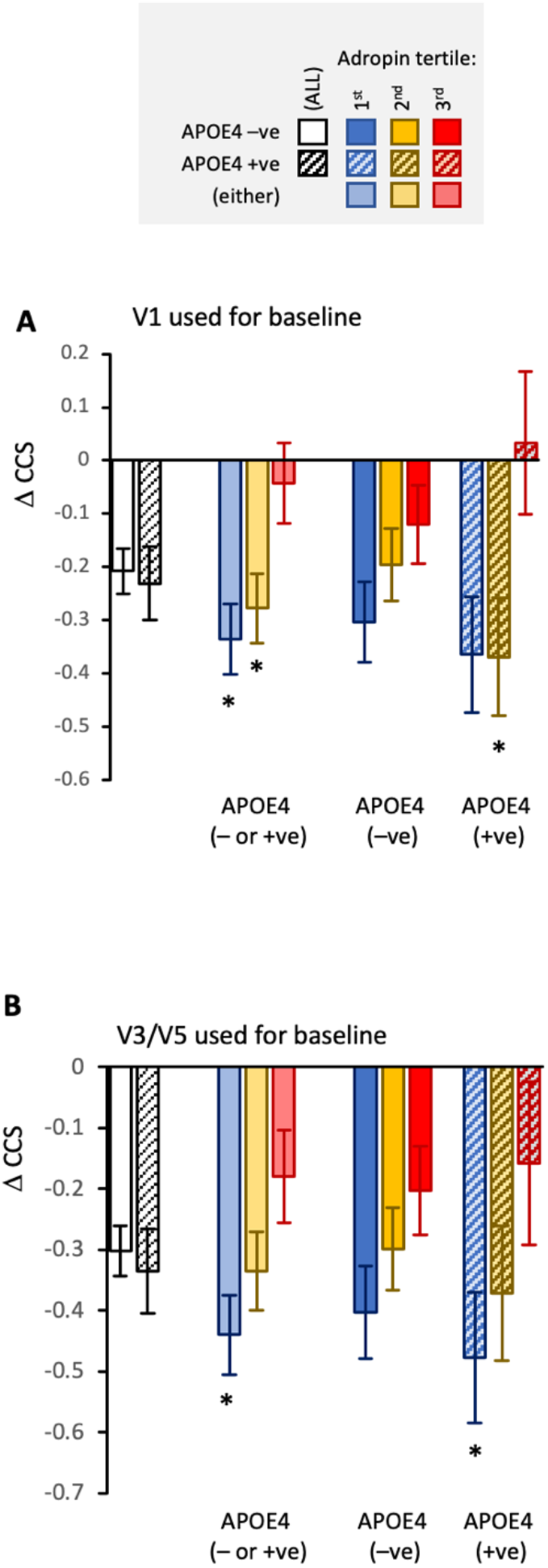
Comparing the differences in ΔCCS between adropin tertile for participants identified by APOE status. The data shown are estimated marginal means and SEM adjusted for age, years between baseline and final measurement, education, and medications. For data using V1 as baseline (**A**), n=123, 140, and 119 (APOE4 either negative or positive); n=95, 105, 100 (APOE4 negative), or n=42,41, and 31 (APOE4 positive). For data using V3/5 as baseline (**B**), n=130, 139, and 112 (APOE4 either negative or positive); n=93, 99, or 87 (APOE4 negative or), or n=41,40, and 26 (APOE4 positive). Significantly different from the 3^rd^ adropin tertile, ^*^P<0.05.

Higher than normal adropin levels thus still correlate with protection from cognitive decline irrespective of APOE genotype.

### Relationships between plasma adropin concentrations and neurodegenerative markers

RNA-seq data indicate that ENHO expression correlates positively with Aβ protein levels in post-mortem brain samples from people of advanced age (30). Low normalized plasma Aβ_42/40_ ratio correlate with higher levels of cortical Aβ and increased risk of AD (7). Plasma adropin concentrations correlated positively with normalized Aβ_42/40_ ratio (ρ=0.101, P<0.05, n=422). The normalized plasma Aβ_42/40_ ratio also differed between adropin tertiles (F_2,422_=4.502, P=0.012), with higher ratios in the 3^rd^ adropin tertile (**Table 2**). Removal of a single outlier (male in the 3^rd^ adropin tertile with an Aβ_42/40_ ratio of 0.3341, 12 SD from the mean) did not affect the correlation (ρ=0.100, P<0.05, n=421). However, difference between adropin tertiles was now a trend (F_2,421_=2.709, P=0.068). Removal of the outlier reduced the estimated marginal mean for the 3^rd^ adropin tertile (0.115±0.014 for both sexes, n=135; 0.114±0.012 for males, n=53).

Blood levels of NfL are a biomarker of neuroaxonal damage that occurs with inflammation, neurodegeneration, and cerebrovascular diseases (6). There was a significant interaction between sex and adropin tertile (F_2,419_=3.396, P=0.034). Plasma NfL concentrations were elevated in males in the 1^st^ adropin tertile when compared to the 2^nd^ and 3^rd^ adropin tertiles, and when compared to females in the 1^st^ adropin tertile (**Table 2**).

The APOE4 allele is the strongest genetic risk factor identified for AD (41, 42). In the participants used for this study, the distribution of carriers between adropin tertiles was not significantly different (**Table 2**).

Circulating levels of inflammatory markers (C-reactive protein, interleukin-6, soluble tumor necrosis factor receptor-1, monocyte chemoattractant protein-1, and growth/differentiation factor-15) have been linked to declining mental and physical capacity in the MAPT study (43). However, we found no consistent pattern between tertiles in the levels of other systemic markers of inflammatory conditions and cellular stress between adropin tertiles. (**Supplemental Table S1**).

### Cross-section correlation between adropin levels and cardiovascular disease risk factors

Adropin has been linked to vascular function (23-26). Simple modelling using heart rate and blood pressure measurement recorded on V3 and V5 indicated positive correlations between plasma adropin concentrations and heart rate (standing ρ=0.159, P<0.001, N=448; prone position ρ=0.105, P<0.05, N=447). Heart rate and diastolic blood pressure measurement adjusted for age and BMI were significantly lower in the 1^st^ adropin tertile relative to the 2^nd^ and 3^rd^ tertiles (**Table 3**).

Circulating adropin levels correlate with indices of metabolic homeostasis, including risk factors of cardiovascular disease and diabetes (13, 14). Fasting blood glucose levels were not significantly different, although incident diabetes (fasting glucose >126 mg/dL on V3 or V5) appeared to be more common in the 3^rd^ adropin tertile (**Table 4**). Plasma levels of total and HDL-cholesterol were significantly lower in the 1^st^ adropin tertile relative to the 2^nd^ and 3^rd^ adropin tertiles (**Table 5**). However, plasma concentrations of ApoA1 (a component of HDL) while correlating HDL-cholesterol levels (ρ=0.562, P<0.001), were not significantly different be tertile (**Table 5**). High plasma ApoB levels are a strong predictor of cardiovascular risk (44). For these participants, there was an interaction between sex and adropin tertile. Plasma ApoB100 concentrations were lower in the 1^st^ adropin tertile in males relative to the other tertile, but there were no differences in females (**Table 5**). Fasting triglyceride concentrations were not significantly different between tertiles (**Table 5**). Finally, prescriptions for glucose and lipid lowering drugs were also similar between tertiles (**Tables 4, 5**).

**Table 4.**
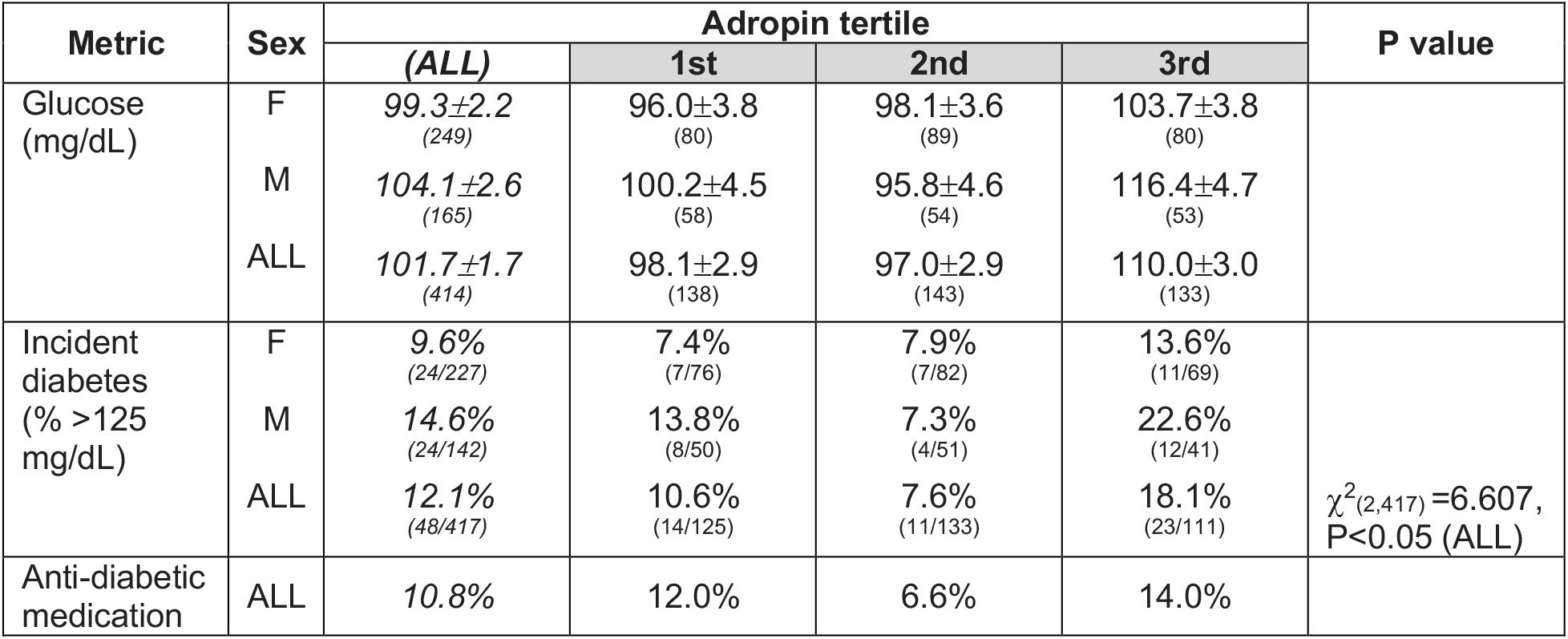
Cross-sectional analysis of indices of glucose and lipid homeostasis (age, BMI adjusted) between adropin tertile. The values shown were measured during the same visit (V3, V5); the percent reporting medications during the first 3 years of the study for treating diabetes and dyslipidemia are shown. P values for covariates less than 0.05 are shown in *italics*.

| Metric | Sex | Adiponin tertile |  |  |  | P value |
| --- | --- | --- | --- | --- | --- | --- |
|  |  | (ALL) | 1st | 2nd | 3rd |  |
| Glucose (mg/dL) | F | 99.3±2.2<br>(249) | 96.0±3.8<br>(80) | 98.1±3.6<br>(89) | 103.7±3.8<br>(80) |  |
|  | M | 104.1±2.6<br>(165) | 100.2±4.5<br>(58) | 95.8±4.6<br>(54) | 116.4±4.7<br>(53) |  |
|  | ALL | 101.7±1.7<br>(414) | 98.1±2.9<br>(138) | 97.0±2.9<br>(143) | 110.0±3.0<br>(133) |  |
| Incident diabetes (% >125 mg/dL) | F | 9.6%<br>(24/227) | 7.4%<br>(7/76) | 7.9%<br>(7/82) | 13.6%<br>(11/69) | $\chi^2_{(2,417)}=6.607$,<br>P<0.05 (ALL) |
|  | M | 14.6%<br>(24/142) | 13.8%<br>(8/50) | 7.3%<br>(4/51) | 22.6%<br>(12/41) |  |
|  | ALL | 12.1%<br>(48/417) | 10.6%<br>(14/125) | 7.6%<br>(11/133) | 18.1%<br>(23/111) |  |
| Anti-diabetic medication | ALL | 10.8% | 12.0% | 6.6% | 14.0% |  |

**Table 5.** Cross-sectional analysis of indices of lipid homeostasis adjusted for age and BMI between adropin tertile. The values shown were measured during the same visit (V3, V5); the percent reporting medications during the first 3 years of the study for treating diabetes and dyslipidemia are shown. P values for covariates less than 0.05 are shown in *italics*.

| Metric | Sex | Adropin tertile |  |  |  | P value |
| --- | --- | --- | --- | --- | --- | --- |
|  |  | (ALL) | 1st | 2nd | 3rd |  |
| Cholesterol (mg/dL) | F | 226.8±2.6<br>(250) | 224.8±4.5<br>(81) | 230.4±4.3<br>(90) | 225.2±4.6<br>(79) | *Age, P<0.05* |
|  | M | 205.7±3.2<br>(164) | 192.5±5.3<sup>1</sup><br>(58) | 212.3±5.5<br>(54) | 212.4±5.6<br>(52) | Sex, P<0.001 |
|  | ALL | 216.3±2.0<br>(414) | 208.7±3.5<sup>2</sup><br>(138) | 221.4±3.5<br>(144) | 218.8±3.6<br>(132) | Tertile, P<0.05 |
| HDL-cholesterol (mg/dL) | F | 70.2±0.9<br>(249) | 64.8±1.7<sup>3</sup><br>(80) | 72.1±1.6<br>(90) | 73.9±1.7<br>(79) | *BMI, P<0.001* |
|  | M | 58.7±1.2<br>(164) | 57.2±2.0<br>(58) | 58.5±2.0<br>(54) | 60.4±2.1<br>(52) | Sex, P<0.001 |
|  | ALL | 64.5±0.7<br>(413) | 61.0±1.3<sup>4</sup><br>(138) | 65.3±1.3<br>(144) | 67.1±1.3<br>(132) | Tertile, P<0.005 |
| ApoA1 (mg/dL) | F | 135.4±2.1<br>(240) | 130.7±3.9<br>(67) | 137.7±3.5<br>(85) | 137.7±3.4<br>(88) | *BMI, P<0.001* |
|  | M | 122.7±2.5<br>(167) | 116.9±4.4<br>(53) | 127.6±4.2<br>(56) | 123.5±4.2<br>(58) | Sex, P<0.001 |
|  | ALL | 129.0±1.6<br>(407) | 123.8±2.9<br>(120) | 132.6±2.7<br>(139) | 130.6±2.7<br>(140) |  |
| ApoB100 (mg/dL) | F | 87.3±1.8<br>(240) | 85.0±3.4<br>(67) | 88.7±3.0<br>(85) | 88.1±3.0<br>(88) | Tertile, P<0.001 |
|  | M | 83.9±2.1<br>(167) | 71.7±3.8<sup>5</sup><br>(53) | 94.4±3.7<br>(56) | 85.4±3.6<br>(58) | Sex X tertile, P<0.05 |
|  | ALL | 85.6±1.4<br>(407) | 78.4±2.5<sup>6</sup><br>(120) | 91.6±2.4<br>(141) | 86.8±2.4<br>(146) |  |
| Triglycerides (mg/dL) | F | 142.0±4.3<br>(250) | 154.9±7.6<br>(80) | 141.6±7.2<br>(90) | 129.5±7.6<br>(80) | *BMI, P<0.001* |
|  | M | 145.4±5.3<br>(164) | 137.7±8.9<br>(58) | 152.0±9.2<br>(54) | 146.6±9.4<br>(52) |  |
|  | ALL | 143.7±3.4<br>(414) | 146.3±5.8<br>(138) | 146.8±5.8<br>(144) | 138.1±6.0<br>(132) |  |
| Lipid-lowering medication | ALL | 44.2% | 49.3% | 44.7% | 38.7% |  |

<sup>1</sup> P<0.05 vs. 2<sup>nd</sup>, 3<sup>rd</sup> adropin tertile;
<sup>2</sup> P<0.05 vs. 2<sup>nd</sup> adropin tertile;
<sup>3</sup> P ≤0.005 vs. 2<sup>nd</sup>, 3<sup>rd</sup> adropin tertile;
<sup>4</sup> P<0.005 vs. 3<sup>rd</sup> adropin tertile;
<sup>5</sup> P<0.05 within column (between sex), P<0.001 vs. 2<sup>nd</sup> adropin tertile, P<0.05 vs. 3<sup>rd</sup> adropin tertile
<sup>6</sup> P<0.001 vs. 2<sup>nd</sup> adropin tertile, P<0.05 vs. 3<sup>rd</sup> adropin tertile

## DISCUSSION

The major findings of this study suggest that plasma adropin concentrations are a new blood-based biomarker of risk for cognitive decline. The results from mouse studies suggest that plasma adropin concentrations could be a modifiable risk factor. Increasing adropin activity in male C57BL/6J mice aged 18-20 months, corresponding to a human age of 56-69 years, improves cognitive ability as assessed by tests of spatial learning and memory (30).

The main outcomes from the current study suggest that people of advanced age with higher levels of adropin in the circulation are protected from cognitive decline. This interpretation is consistent with data from experiments using mouse models that demonstrated increasing adropin activity enhances cognitive ability (28-30, 33). This finding is important for two reasons. First, it suggests an opportunity for developing an additional plasma biomarker indicating risk of cognitive decline. Secondly, results from preclinical experiments using mouse models indicate the people with lower-than-normal circulating adropin levels might benefit from therapies that increase adropin signaling. Knowledge of the source and sequences of the adropin peptides will be critical for the development of adropin analogs for use in clinical studies.

Data from The Aging, Dementia and TBI Study indicated relationships between expression of the ENHO transcript encoding adropin and transcriptomic and protein signatures of brain health that could be interpreted in two ways (30). On the one hand, there was a positive correlation with transcriptomic signatures of mitochondrial function and synaptic plasticity. High adropin activity in the brain could therefore correlate positively with brain energy metabolism and neural activity. However, positive associations with protein markers of A? accumulation and Tau pathology in the brain were also noted. While the results from mouse studies suggest the relationship between adropin and cognitive health would be positive as opposed to negative, either outcome was considered possible for the current study.

The relationships between plasma adropin levels and either normalized plasma Aβ_42/40_ ratio or plasma NfL concentrations are also an interesting observation. Both results are consistent with higher-than-normal circulating adropin levels indicating neuroprotection in the context of aging. Higher plasma Aβ_42/40_ ratios are an indicator of reduced risk of amyloidosis, cognitive decline, or developing AD dementia (7). The higher levels in participants in the 3^rd^ adropin tertile is therefore consistent with delayed neurodegeneration which could contribute to protection from cognitive decline. For NfL, there appears to be sexual dimorphism with higher levels observed in participants with lower-than-normal plasma adropin concentrations. On the other hand, there was no relationship between plasma adropin concentrations and systemic markers of inflammation. This suggests that plasma adropin concentrations are more closely correlated to neurodegenerative conditions, as opposed to inflammation. Clearly, further studies examining the relationship between circulating adropin levels and neurodegeneration are warranted. Indeed, one of the limitations of the current study is the study population, who were selected for expressing spontaneous memory complaint and/or other signs of frailty. Of the group studied here, all but one had low plasma Aβ_42/40_ ratios. It would be of interest to extend the study to include people of advanced age with a broader spectrum of cognitive abilities and neuropathology.

A significant caveat to interpreting the results from this study is that the lack of information on the source of adropin peptide in the circulation. In mice, both protein and mRNA expression of adropin are high in the brain relative to other tissues (9, 10). In nonhuman primates and humans, expression of the mRNA encoding adropin is far higher in the nervous system relative to other tissues (30, 31). Whether circulating adropin levels correlate with expression in the nervous system, or with levels in cerebrospinal fluid, is not known and needs to be studied.

The interpretation of this data is that circulating adropin levels correlate with activity in the nervous system is one possible interpretation. However, experiments using mouse models of ischemia indicate the blood-brain barrier is a critical target for adropin (28, 29). The blood-brain barrier has a critical role in maintaining brain health and preventing dementia during aging (45, 46). Further experiments are needed to determine whether circulating adropin levels correlate directly with activity of the peptide in the brain or indicate an indirect relationship reflecting the relative health of the blood brain barrier. Advances in our knowledge of the signaling mechanisms involved are also required. The only signaling pathway currently known to be necessary for adropin activity in the brain involves eNOS signaling (29).

The current study also compared plasma adropin concentrations with indicators of cardiovascular and metabolic homeostasis. In a cross-sectional analysis of the data collected at V3 and V5, participants with high adropin appear to be more at risk for cardiovascular disease, indicated by higher levels of total cholesterol and ApoB100. This result is starkly different from studies comparing plasma adropin concentrations with indices of cholesterol in younger people which showed the opposite but in males only (47). It is possible that relationships between plasma adropin concentration and indices of cholesterol metabolism are age specific. Another possible explanation involves differences in the environments experienced by populations living in the United States and France.

In the current study, people with lower-than-normal circulating adropin concentrations also had a relatively low heart rate. There was also trend for an interaction between sex and adropin tertile for diastolic blood pressure. These observations could indicate an inverse relationship between circulating adropin levels and vascular condition in this population. The increased relative risk of cognitive decline with lower-than-normal plasma adropin concentrations thus does not correlate with cardiovascular risk factors in this study.

The variables explaining differences in plasma adropin concentrations between people are not clearly defined. However, increased adropin expression and/or release could be a protective-adaptive response to stress. Indeed, experiments in mice indicate that increased adropin expression in non-neural tissues is a component of a cellular-stress response (17, 48). In addition, in humans the development of insulin resistance and dyslipidemia due to the consumption of fructose beverages associates with an increase in plasma adropin concentrations (49). On the other hand, there was no correlation with circulating markers of axonal damage, inflammation or cellular stress response in this study. Moreover, there was no difference in the prescription of medications against diabetes, dyslipidemia, or diabetes between adropin tertile. The relationships observed between plasma adropin concentrations and indices of cardiovascular health and lipid metabolism thus reflect the conditions in each of the participants on the day of measurement. The relationship between circulating adropin and risk of cognitive decline thus appears to involve other pathways that require further study.

In conclusion, the results of this study indicate that high plasma adropin concentrations associate with attenuated cognitive decline in older people. Measurement of circulating adropin levels could have added value in that modification through enhancement of adropin signaling using synthetic protein- or genetic-approaches could be possible (28, 30, 33). Further studies exploring the relationships between circulating adropin and neurological aging, and the mechanisms that underly this association, are clearly warranted.

## Data Availability

A de-identified data set can be made available to academic researchers who provide a methodologically sound proposal that is approved by the MAPT/DSA group. Enquiries should be directed to.

## Acknowledgments

AAB and ADN acknowledge financial support provided by Saint Louis University. The authors thank Prof. Randall J. Bateman (Department of Neurology, Washington University School of Medicine, 660 South Euclid Avenue, Box 8111, St. Louis, MO 63110 USA) for measurement of plasma A?42/40 ratio. The authors also thanks Dr. Theodore Malmstrom (Department of Psychiatry & Behavioiral Neuroscience, Saint Louis University School of Medicine, 1438 South Grand Boulevard, St. Louis, MO 63104 USA) for discussion on statistical analysis. The MAPT study was supported by grants from the Gérontopôle of Toulouse, the French Ministry of Health (PHRC 2008, 2009), Pierre Fabre Research Institute (manufacturer of the omega-3 supplement), ExonHit Therapeutics SA, and Avid Radiopharmaceuticals Inc. The promotion of this study was supported by the University Hospital Center of Toulouse. The data sharing activity was supported by the Association Monegasque pour la Recherche sur la maladie d’Alzheimer (AMPA) and the INSERM-University of Toulouse III UMR 1295 (CERPOP) Research Unit

## Contributors

AAB, ADN, BV, and JEM co-designed the study. GA, AAB, and ADN collected the data. GA verified the underlying data. AAB analyzed the data and prepared the first draft of the manuscript. All authors contributed to the interpretation of the results and critically reviewed the manuscript. All authors read and approved the final version of the manuscript.

## Ethics declarations

## Ethical approval

The MAPT was approved by the ethics committee in Toulouse (CPP SOOM II). All research was performed in accordance with relevant guidelines/regulations.

## Informed consent

Written consent forms were obtained from all participants.

## Conflict of Interest

The authors declare no competing interests.

## Collaborators (members of the MAPTDSA group)

*Principal investigator*: Bruno Vellas (Toulouse); *Coordination*: Sophie Guyonnet ; *Project leader*: Isabelle Carrié ; CRA: Lauréane Brigitte ; *Investigators*: Catherine Faisant, Françoise Lala, Julien Delrieu, Hélène Villars ; *Psychologists*: Emeline Combrouze, Carole Badufle, Audrey Zueras ; *Methodology, statistical analysis and data management*: Sandrine Andrieu, Christelle Cantet, Christophe Morin; *Multidomain group*: Gabor Abellan Van Kan, Charlotte Dupuy, Yves Rolland (physical and nutritional components), Céline Caillaud, Pierre-Jean Ousset (cognitive component), Françoise Lala (preventive consultation). The cognitive component was designed in collaboration with Sherry Willis from the University of Seattle, and Sylvie Belleville, Brigitte Gilbert and Francine Fontaine from the University of Montreal. *Co-Investigators in associated centres*: Jean-François Dartigues, Isabelle Marcet, Fleur Delva, Alexandra Foubert, Sandrine Cerda (Bordeaux); Marie-Noëlle-Cuffi, Corinne Costes (Castres); Olivier Rouaud, Patrick Manckoundia, Valérie Quipourt, Sophie Marilier, Evelyne Franon (Dijon); Lawrence Bories, Marie-Laure Pader, Marie-France Basset, Bruno Lapoujade, Valérie Faure, Michael Li Yung Tong, Christine Malick-Loiseau, Evelyne Cazaban-Campistron (Foix); Françoise Desclaux, Colette Blatge (Lavaur); Thierry Dantoine, Cécile Laubarie-Mouret, Isabelle Saulnier, Jean-Pierre Clément, Marie-Agnès Picat, Laurence Bernard-Bourzeix, Stéphanie Willebois, Iléana Désormais, Noëlle Cardinaud (Limoges); Marc Bonnefoy, Pierre Livet, Pascale Rebaudet, Claire Gédéon, Catherine Burdet, Flavien Terracol (Lyon), Alain Pesce, Stéphanie Roth, Sylvie Chaillou, Sandrine Louchart (Monaco); Kristel Sudres, Nicolas Lebrun, Nadège Barro-Belaygues (Montauban); Jacques Touchon, Karim Bennys, Audrey Gabelle, Aurélia Romano, Lynda Touati, Cécilia Marelli, Cécile Pays (Montpellier); Philippe Robert, Franck Le Duff, Claire Gervais, Sébastien Gonfrier (Nice); Yannick Gasnier and Serge Bordes, Danièle Begorre, Christian Carpuat, Khaled Khales, Jean-François Lefebvre, Samira Misbah El Idrissi, Pierre Skolil, Jean-Pierre Salles (Tarbes). *MRI group*: Carole Dufouil (Bordeaux), Stéphane Lehéricy, Marie Chupin, Jean-François Mangin, Ali Bouhayia (Paris); Michèle Allard (Bordeaux); Frédéric Ricolfi (Dijon); Dominique Dubois (Foix); Marie Paule Bonceour Martel (Limoges); François Cotton (Lyon); Alain Bonafé (Montpellier); Stéphane Chanalet (Nice); Françoise Hugon (Tarbes); Fabrice Bonneville, Christophe Cognard, François Chollet (Toulouse). *PET scans group*: Pierre Payoux, Thierry Voisin, Julien Delrieu, Sophie Peiffer, Anne Hitzel, (Toulouse); Michèle Allard (Bordeaux); Michel Zanca (Montpellier); Jacques Monteil (Limoges); Jacques Darcourt (Nice). Medico-economics group: Laurent Molinier, Hélène Derumeaux, Nadège Costa (Toulouse). *Biological sample collection*: Bertrand Perret, Claire Vinel, Sylvie Caspar-Bauguil (Toulouse). *Safety management*: Pascale Olivier-Abbal. *DSA Group*: Sandrine Andrieu, Christelle Cantet, Nicola Coley

---

**Supplemental Table 1.** Plasma concentrations of inflammatory markers (soluble tumor necrosis factor receptor, sTNFR-1; monocyte chemoattractant protein 1, MCP-1; interleukin 6, IL-6, C-reactive protein, CRP) and cellular stress (plasma growth differentiation factor 15, GDF-15). The data shown were measured at V3 and V5 and are presented as estimated marginal means±SE adjusted for age. P values for covariates less than 0.05 are shown in *italics*.

| Metric | Sex | Adropin tertile |  |  |  | P value |
| --- | --- | --- | --- | --- | --- | --- |
|  |  | (ALL) | 1st | 2nd | 3rd |  |
| sTNFR-1 (pg/mL) | F | 1334±31<br>(240) | 1283±57<br>(68) | 1374±52<br>(84) | 1345±50<br>(88) | *Age, P<0.001*<br>*Sex, P<0.05* |
| | M | 1447±36<br>(167) | 1493±65<br>(52) | 1428±62<br>(57) | 1418±62<br>(58) | |
| | ALL | 1390±24<br>(407) | 1388±43<br>(120) | 1401±40<br>(141) | 1382±40<br>(146) | |
| MCP-1 (pg/mL) | F | 248±6<br>(240) | 244±11<br>(64) | 236±10<br>(84) | 265±10<br>(88) | |
| | M | 242±7<br>(167) | 235±13<br>(52) | 229±12<br>(57) | 262±12<br>(58) | |
| | ALL | 245±5<br>(407) | 239±9<br>(120) | 233±8<br>(141) | 263±8<br>(146) | |
| IL-6 (pg/mL) | F | 5.65±1.19<br>(240) | 9.36±2.23<br>(68) | 4.02±2.01<br>(84) | 3.57±1.96<br>(88) | |
| | M | 3.97±1.42<br>(167) | 3.90±2.55<br>(52) | 3.69±2.43<br>(57) | 4.31±2.42<br>(58) | |
| | ALL | 4.81±0.93<br>(407) | 6.63±1.69<br>(120) | 3.86±1.58<br>(141) | 3.94±1.56<br>(146) | |
| CRP (mg/L) | F | 3.07±0.32<br>(245) | 2.75±0.55<br>(80) | 3.50±0.53<br>(87) | 2.96±0.56<br>(78) | |
| | M | 2.93±0.39<br>(163) | 2.30±0.66<br>(56) | 2.87±0.67<br>(55) | 3.62±0.69<br>(52) | |
| | ALL | 3.00±0.25<br>(408) | 2.52±0.43<br>(136) | 3.18±0.43<br>(142) | 3.29±0.44<br>(130) | |
| GDF-15 (pg/mL) | F | 1198±32<br>(239) | 1225±59<br>(68) | 1183±54<br>(83) | 1184±52<br>(88) | *Age, P<0.001*<br>*Sex, P<0.001* |
| | M | 1438±38<br>(167) | 1571±68<br>(52) | 1382±64<br>(57) | 1361±64<br>(58) | |
| | ALL | 1318±25<br>(406) | 1398±45<br>(120) | 1283±42<br>(140) | 1273±41<br>(146) | |

## Notes

### Competing Interest Statement

The authors have declared no competing interest.

### Author Declarations

The MAPT was approved by the ethics committee in Toulouse (CPP SOOM II). Written consent forms were obtained from all participants. All research was performed in accordance with relevant guidelines/regulations.The MAPT Study was registered at ClinicalTrials.gov [no.: NCT00672685], approved by the French Ethical Committee located in Toulouse (CPP SOOM II) and authorized by the French Health Authority.

### Summary of Updates

Change of title; Edited Abstract and Introduction; Updated statistical analysis; revision of Figure 1 (split into 3 figures).

## REFERENCES

1. 2023 Alzheimer’s disease facts and figures. Alzheimers Dement. 2023;19(4):1598–695.

2. Lutz W, Sanderson W, Scherbov S. The coming acceleration of global population ageing. Nature. 2008;451(7179):716–9.

3. Collaborators GBDDF. Estimation of the global prevalence of dementia in 2019 and forecasted prevalence in 2050: an analysis for the Global Burden of Disease Study 2019. Lancet Public Health. 2022;7(2):e105–e25.

4. Jack CR, Jr., Bennett DA, Blennow K, Carrillo MC, Dunn B, Haeberlein SB, et al. NIA-AA Research Framework: Toward a biological definition of Alzheimer’s disease. Alzheimers Dement. 2018;14(4):535–62.

5. Teunissen CE, Verberk IMW, Thijssen EH, Vermunt L, Hansson O, Zetterberg H, et al. Blood-based biomarkers for Alzheimer’s disease: towards clinical implementation. Lancet Neurol. 2022;21(1):66–77.

6. Abu-Rumeileh S, Abdelhak A, Foschi M, D’Anna L, Russo M, Steinacker P, et al. The multifaceted role of neurofilament light chain protein in non-primary neurological diseases. Brain. 2023;146(2):421–37.

7. Brand AL, Lawler PE, Bollinger JG, Li Y, Schindler SE, Li M, et al. The performance of plasma amyloid beta measurements in identifying amyloid plaques in Alzheimer’s disease: a literature review. Alzheimers Res Ther. 2022;14(1):195.

8. Brockmann R, Nixon J, Love BL, Yunusa I. Impacts of FDA approval and Medicare restriction on antiamyloid therapies for Alzheimer’s disease: patient outcomes, healthcare costs, and drug development. Lancet Reg Health Am. 2023;20:100467.

9. Kumar KG, Trevaskis JL, Lam DD, Sutton GM, Koza RA, Chouljenko VN, et al. Identification of adropin as a secreted factor linking dietary macronutrient intake with energy homeostasis and lipid metabolism. Cell Metab. 2008;8(6):468–81.

10. Wong CM, Wang Y, Lee JT, Huang Z, Wu D, Xu A, et al. Adropin is a brain membrane-bound protein regulating physical activity via the NB-3/Notch signaling pathway in mice. J Biol Chem. 2014;289(37):25976–86.

11. Jumper J, Evans R, Pritzel A, Green T, Figurnov M, Ronneberger O, et al. Highly accurate protein structure prediction with AlphaFold. Nature. 2021;596(7873):583–9.

12. Tusnady GE, Simon I. The HMMTOP transmembrane topology prediction server. Bioinformatics. 2001;17(9):849–50.

13. Butler AA, Havel PJ. Adropin and insulin resistance: Integration of endocrine, circadian, and stress signals regulating glucose metabolism. Obesity (Silver Spring). 2021;29(11):1799–801.

14. Mushala BAS, Scott I. Adropin: a hepatokine modulator of vascular function and cardiac fuel metabolism. Am J Physiol Heart Circ Physiol. 2021;320(1):H238–H44.

15. Barzilai N, Huffman DM, Muzumdar RH, Bartke A. The critical role of metabolic pathways in aging. Diabetes. 2012;61(6):1315–22.

16. Gao S, Ghoshal S, Zhang L, Stevens JR, McCommis KS, Finck BN, et al. The peptide hormone adropin regulates signal transduction pathways controlling hepatic glucose metabolism in a mouse model of diet-induced obesity. J Biol Chem. 2019;294(36):13366–77.

17. Banerjee S, Ghoshal S, Stevens JR, McCommis KS, Gao S, Castro-Sepulveda M, et al. Hepatocyte expression of the micropeptide adropin regulates the liver fasting response and is enhanced by caloric restriction. J Biol Chem. 2020;295(40):13753–68.

18. Thapa D, Xie B, Manning JR, Zhang M, Stoner MW, Huckestein BR, et al. Adropin reduces blood glucose levels in mice by limiting hepatic glucose production. Physiol Rep. 2019;7(8):e14043.

19. Gao S, McMillan RP, Zhu Q, Lopaschuk GD, Hulver MW, Butler AA. Therapeutic effects of adropin on glucose tolerance and substrate utilization in diet-induced obese mice with insulin resistance. Mol Metab. 2015;4(4):310–24.

20. Chen X, Chen S, Shen T, Yang W, Chen Q, Zhang P, et al. Adropin regulates hepatic glucose production via PP2A/AMPK pathway in insulin-resistant hepatocytes. FASEB J. 2020.

21. Gao S, McMillan RP, Jacas J, Zhu Q, Li X, Kumar GK, et al. Regulation of substrate oxidation preferences in muscle by the peptide hormone adropin. Diabetes. 2014;63(10):3242–52.

22. Altamimi TR, Gao S, Karwi QG, Fukushima A, Rawat S, Wagg CS, et al. Adropin regulates cardiac energy metabolism and improves cardiac function and efficiency. Metabolism. 2019;98:37–48.

23. Jurrissen TJ, Ramirez-Perez FI, Cabral-Amador FJ, Soares RN, Pettit-Mee RJ, Betancourt-Cortes EE, et al. Role of adropin in arterial stiffening associated with obesity and type 2 diabetes. Am J Physiol Heart Circ Physiol. 2022;323(5):H879–H91.

24. Fujie S, Hasegawa N, Horii N, Uchida M, Sanada K, Hamaoka T, et al. Aerobic Exercise Restores Aging-Associated Reductions in Arterial Adropin Levels and Improves Adropin-Induced Nitric Oxide-Dependent Vasorelaxation. J Am Heart Assoc. 2021:e020641.

25. Fujie S, Hasegawa N, Kurihara T, Sanada K, Hamaoka T, Iemitsu M. Association between aerobic exercise training effects of serum adropin level, arterial stiffness, and adiposity in obese elderly adults. Appl Physiol Nutr Metab. 2017;42(1):8–14.

26. Fujie S, Hasegawa N, Sato K, Fujita S, Sanada K, Hamaoka T, et al. Aerobic exercise training-induced changes in serum adropin level are associated with reduced arterial stiffness in middle-aged and older adults. Am J Physiol Heart Circ Physiol. 2015;309(10):H1642–7.

27. Gunraj RE, Yang C, Liu L, Larochelle J, Candelario-Jalil E. Protective roles of adropin in neurological disease. Am J Physiol Cell Physiol. 2023;324(3):C674–C8.

28. Yang C, Liu L, Lavayen BP, Larochelle J, Gunraj RE, Butler AA, et al. Therapeutic Benefits of Adropin in Aged Mice After Transient Ischemic Stroke via Reduction of Blood-Brain Barrier Damage. Stroke. 2023;54(1):234–44.

29. Yang C, Lavayen BP, Liu L, Sanz BD, DeMars KM, Larochelle J, et al. Neurovascular protection by adropin in experimental ischemic stroke through an endothelial nitric oxide synthase-dependent mechanism. Redox Biol. 2021;48:102197.

30. Banerjee S, Ghoshal S, Girardet C, DeMars KM, Yang C, Niehoff ML, et al. Adropin correlates with aging-related neuropathology in humans and improves cognitive function in aging mice. NPJ Aging Mech Dis. 2021;7(1):23.

31. Butler AA, Zhang J, Price CA, Stevens JR, Graham JL, Stanhope KL, et al. Low plasma adropin concentrations increase risks of weight gain and metabolic dysregulation in response to a high-sugar diet in male nonhuman primates. J Biol Chem. 2019;294(25):9706–19.

32. Yang C, DeMars KM, Candelario-Jalil E. Age-Dependent Decrease in Adropin is Associated with Reduced Levels of Endothelial Nitric Oxide Synthase and Increased Oxidative Stress in the Rat Brain. Aging Dis. 2018;9(2):322–30.

33. Ghoshal S, Banerjee S, Zhang J, Niehoff ML, Farr SA, Butler AA. Adropin transgenesis improves recognition memory in diet-induced obese LDLR-deficient C57BL/6J mice. Peptides. 2021;146:170678.

34. Vellas B, Carrie I, Gillette-Guyonnet S, Touchon J, Dantoine T, Dartigues JF, et al. Mapt Study: A Multidomain Approach for Preventing Alzheimer’s Disease: Design and Baseline Data. J Prev Alzheimers Dis. 2014;1(1):13–22.

35. Andrieu S, Guyonnet S, Coley N, Cantet C, Bonnefoy M, Bordes S, et al. Effect of long-term omega 3 polyunsaturated fatty acid supplementation with or without multidomain intervention on cognitive function in elderly adults with memory complaints (MAPT): a randomised, placebo-controlled trial. Lancet Neurol. 2017;16(5):377–89.

36. Giudici KV, Barreto PS, Guyonnet S, Morley JE, Nguyen AD, Aggarwal G, et al. TNFR-1 and GDF-15 are associated with plasma neurofilament light chain and progranulin among community-dwelling older adults: a secondary analysis of the MAPT Study. J Gerontol A Biol Sci Med Sci. 2022.

37. He L, de Souto Barreto P, Aggarwal G, Nguyen AD, Morley JE, Li Y, et al. Plasma Abeta and neurofilament light chain are associated with cognitive and physical function decline in non-dementia older adults. Alzheimers Res Ther. 2020;12(1):128.

38. Lu WH, Giudici KV, Rolland Y, Guyonnet S, Li Y, Bateman RJ, et al. Prospective Associations between Plasma Amyloid-Beta 42/40 and Frailty in Community-Dwelling Older Adults. J Prev Alzheimers Dis. 2021;8(1):41–7.

39. Giudici KV, de Souto Barreto P, Guyonnet S, Li Y, Bateman RJ, Vellas B, et al. Assessment of Plasma Amyloid-beta42/40 and Cognitive Decline Among Community-Dwelling Older Adults. JAMA Netw Open. 2020;3(12):e2028634.

40. Coley N, Gallini A, Ousset PJ, Vellas B, Andrieu S, GuidAge study g. Evaluating the clinical relevance of a cognitive composite outcome measure: An analysis of 1414 participants from the 5-year GuidAge Alzheimer’s prevention trial. Alzheimers Dement. 2016;12(12):1216–25.

41. Carlstrom K, Castelo-Branco G. Alzheimer’s risk variant APOE4 linked to myelin-assembly malfunction. Nature. 2022;611(7937):670–1.

42. Dolgin E. This is how an Alzheimer’s gene ravages the brain. Nature. 2022;611(7937):649.

43. Lu WH, Gonzalez-Bautista E, Guyonnet S, Lucas A, Parini A, Walston JD, et al. Plasma inflammation-related biomarkers are associated with intrinsic capacity in community-dwelling older adults. J Cachexia Sarcopenia Muscle. 2023;14(2):930–9.

44. Glavinovic T, Thanassoulis G, de Graaf J, Couture P, Hegele RA, Sniderman AD. Physiological Bases for the Superiority of Apolipoprotein B Over Low-Density Lipoprotein Cholesterol and Non-High-Density Lipoprotein Cholesterol as a Marker of Cardiovascular Risk. J Am Heart Assoc. 2022;11(20):e025858.

45. Banks WA, Reed MJ, Logsdon AF, Rhea EM, Erickson MA. Healthy aging and the blood-brain barrier. Nat Aging. 2021;1(3):243–54.

46. Segarra M, Aburto MR, Acker-Palmer A. Blood-Brain Barrier Dynamics to Maintain Brain Homeostasis. Trends Neurosci. 2021;44(5):393–405.

47. Ghoshal S, Stevens JR, Billon C, Girardet C, Sitaula S, Leon AS, et al. Adropin: An endocrine link between the biological clock and cholesterol homeostasis. Mol Metab. 2018;8:51–64.

48. Chen X, Xue H, Fang W, Chen K, Chen S, Yang W, et al. Adropin protects against liver injury in nonalcoholic steatohepatitis via the Nrf2 mediated antioxidant capacity. Redox Biol. 2019;21:101068.

49. Butler AA, St-Onge MP, Siebert EA, Medici V, Stanhope KL, Havel PJ. Differential Responses of Plasma Adropin Concentrations To Dietary Glucose or Fructose Consumption In Humans. Sci Rep. 2015;5:14691.

